# Understanding Economic and Health Factors Impacting the Spread of COVID-19 Disease

**DOI:** 10.1101/2020.04.10.20058222

**Authors:** Aleksandr Farseev, Yu-Yi Chu-Farseeva, Qi Yang, Daron Benjamin Loo

## Abstract

**Background:** The rapid spread of the Coronavirus 2019 disease (COVID-19) had drastically impacted life all over the world. While some economies are actively recovering from this pestilence, others are experiencing fast and consistent disease spread, compelling governments to impose social distancing measures that have put a halt on routines, especially in densely populated areas.

**Objective:** Aiming at bringing more light on key economic and population health factors affecting the disease spread, this initial study utilizes a quantitative statistical analysis based on the most recent publicly available COVID-19 datasets.

**Methods:** We have applied Pearson Correlation Analysis and Clustering Analysis (X-Means Clustering) techniques on the data obtained by combining multiple datasets related to country economics, medical system & health, and COVID-19 - related statistics. The resulting dataset consisted of COVID-19 Case and Mortality Rates, Economic Statistics, and Population Public Health Statistics for 165 countries reported between 22 January 2020 and 28 March 2020. The correlation analysis was conducted with the significance level α of 0.05. The clustering analysis was guided by the value of Bayesian Information Criterion (BIC) with the bin value b = 1.0 and the cutoff factor c = 0.5, and have provided a stable split into four country-level clusters.

**Results:** The study showed and explained multiple significant relationships between the COVID-19 data and other country-level statistics. We also identified and statistically profiled four major country-level clusters with relation to different aspects of COVID-19 development and country-level economic and health indicators. Specifically, this study identified potential COVID-19 under-reporting traits, as well as various economic factors that impact COVID-19 Diagnosis, Reporting, and Treatment. Based on the country clusters, we also described the four disease development scenarios, which are tightly knit to country-level economic and population health factors. Finally, we highlighted the potential limitation of reporting and measuring COVID-19 and provided recommendations on further in-depth quantitative research.

**Conclusions:** In this study, we first identified possible COVID-19 reporting issues and biases across different countries and regions. Second, we identified crucial factors affecting the speed of COVID-19 disease spread and provided recommendations on choosing and operating economic and health system factors when analyzing COVID-19 progression. Particularly, we discovered that the political system and compliance with international disease control norms are crucial for effective COVID-19 pandemic cessation. However, the role of some widely-adopted measures, such as GHS Health Index, might have been overestimated in lieu of multiple biases and underreporting challenges. Third, we benchmarked our findings against the widely-adopted Global Health Security (GHS) model and found that the latter might be redundant when measuring and forecasting COVID-19 spread, while its individual components could potentially serve as stronger COVID-19 indicators. Fourth, we discovered four clusters of countries characterized by different COVID-19 development scenarios, highlighting the differences of the disease reporting and progression in different economic and health system settings. Finally, we provided recommendations on sophisticated measures and research approaches to be implemented for effective outbreak measurements, evaluation and forecasting. We have supported the latter recommendations by a preliminary regression analysis based on the our-collected dataset. We believe that our work would encourage further in-depth quantitative research along the direction as well as would be of support to public policy development when addressing the COVID-19 crisis worldwide.

## Introduction

The rapid spread of COVID-19 has drastically impacted economies around the world. On 11 March 2020 the disease was officially classified as a pandemic and, as reported on 24 March 2020, it has infected 440,093, and causing 19,748 deaths worldwide, with the highest new case intensities in the USA, Spain, Germany, France, Switzerland, South Korea, United Kingdom (UK), and Hubei Province in China.

In response to such a volatile situation in the world, governments and the scientific communities have been actively studying the underlying principles and possible reasons for the disease spread and progression. For example, Bai et.al. [1] have first discovered that COVID-19 could have been possibly transmitted by asymptomatic carriers, while Wu et.al. [2] conducted a large-scale study based on 72,314 confirmed cases listing important actionable lessons for other societies to apply. Finally, Martin et al. [42] studied the importance of social distancing measures being applied to slow the spread speed of COVID-19 spread pace reduction.

Furthermore, the Computer Science community has analyzed the disease spread from a statistical point of view. Specifically, in [3], the authors witnessed a potential association between COVID-19 mortality rates and health-care resource availability, while Chen et.al. [4] discovered a strong statistical relationship between initial emigration from Wuhan City and the infection spread to other cities in China. Finally, Chinazzi et.al. [5] suggested that travel restrictions to COVID-19 affected areas could be not as effective, as many infected individuals “…have been travelling internationally without being detected…” and as such, sharper restrictive measures are necessary to curb and take control of the outbreak.

Even though significant efforts have been made towards a proper understanding of the COVID-19 outbreak from multiple perspectives, due to the constantly evolving pandemic, emerging new information and data sets, and inaccessibility of public large-scale data, literature based on quantitative research on the outbreak is still relatively sparse. In the study, we hypothesise that the speed of spread of COVID-19 disease is tied to the various economic and health factors, that, in turn, form a country profile as well as reflect the country’s readiness to concur the COVID-19 pandemic. To the best of our knowledge, it is one of the first attempts to build a more holistic view on the COVID-19 development, which hopes to identify and explain relationships between the disease spread and various economic and health factors through quantitative analysis.

## Methods

### Dataset

In this study, we have incorporated the “COVID19 Global Forecasting (Week 2)” dataset [6] that was released by the Kaggle [33] platform. The dataset includes daily updates of the COVID-19 confirmed cases and mortality rates for 173 countries reported by WHO between 22 January 2020 and 28 March 2020. To study the relationships between COVID-19 spread and various economic factors, we merged the original dataset with “Country Statistics - UNData” dataset [7], “Pollution by Country for COVID19 Analysis” dataset [8], and “The World Bank (Demographics)” dataset by cross-matching country names across data sets. We also merged the original dataset with the dataset obtained by parsing the “World Life Expectancy” database [9] website for obtaining information on death rates from different chronic diseases across the world. The selected data indicators were chosen as the key economic and health indicators available in public access aiming at provision of a more holistic view into different country profiles with respect to the COVID-19 pandemic development. Specifically, the Kaggle platform is known to be one of the largest data integrators in the world, where the research community could source the most recent and comprehensive real-time data on the last world-level problems. At the same time, the World life Expectancy datasets are known to be the largest global health and life expectancy databases, allowing for comprehensive statistics about various health factors with respect to different countries. Finally, the UNdata data service was chosen as one of the largest aggregation services for the statistical databases related to country economics, providing us with the economics statistics for most of the countries included in the source Kaggle dataset.

To support this study observation and evaluate our selected indicators against more widely-adopted evaluation system, we have also enriched the dataset by the data provided in Global Health Security Index (GHS) database, which was is claimed to be “…the first comprehensive assessment of global health security capabilities in 195 countries” [41] and, therefore, could serve as an assessment medium for this study. Except for the actual GHS classification, the database also provides indexes measuring various aspects of the health systems:

- **Prevention Index** - Prevention of the emergence or release of pathogens;
- **Detection and Reporting Index** - Early detection and reporting for epidemics of potential international concern;
- **Rapid Response Index** - Rapid response to and mitigation of the spread of an epidemic;
- **Health System Index** - Sufficient and robust health system to treat the sick and protect health workers;
- **Compliance with International Norms Index** - Commitments to improving national capacity, financing plans to address gaps, and adhering to global norms;
- **Risk Environment Index** - Overall risk environment and country vulnerability to biological threats.

After the merging process, the resulting dataset consists of COVID-19 Case and Mortality Rates, Economic Statistics, and Population Health Statistics for 165 countries reported between 22 January 2020 and 28 March 2020. The actual number of the data records in the dataset is 286, as there were COVID-19 statistics in the original data set given for different regions within the same country: 54 regions in the United States of America, 33 regions in China, 10 regions in Canada, 10 regions in France, 8 regions in Australia, 7 regions in the United Kingdom, 4 regions in the Netherlands, and 3 regions in Denmark. Seven countries, namely Bahamas, Congo Brazzaville, Congo Kinshasa, Eswatini, Gambia, Taiwan, and Vietnam were excluded as there were no economics and medical system & health statistics available for them in the merged datasets. A more detailed statistics of the resulting dataset are provided in Table 1. We have also released the dataset for public use [10].

**Table 1:** Detailed Statistics of the COVID-19 Combined Dataset

| Data Indicator Group | Number |
| --- | --- |
| Countries | 165 |
| Regions | 286 |
| Chronic Disease Death Rate Statistics | 32 |
| Age Demographic Groups | 4 |
| Pollution Indicators | 3 |
| Other Economic Factor Statistics | 50 |
| Global Health Security Index (GHS) Indicators | 14 |
| COVID-19 Related Indicators | 2 |
| COVID-19 Confirmed Case Speed Daily Reports | 67 |
| COVID-19 Fatalities Speed Daily Reports | 67 |

### Experimental Setup

As mentioned, the primary objective of this research is to study the relationship between the **speed** of the disease spread and various economic and health factors. Considering the uneven pace of the disease’s geographical spread due to COVID-19’s long incubation period [11], natural migration laws [5] and various government-imposed travel policies [5], it is not feasible to draw the analysis based on the actual daily registered case and fatality rates available in the original Kaggle dataset [6], but rather necessary to perform an additional data pre-processing aiming at establishing holistic data characteristics reflecting the general worldwide COVID-19 spread tendencies. Keeping this in mind, we have performed the following data pre-processing steps:

**Dataset Combination:** Original Dataset [6] was joined by performing Country matching to four auxiliary data sets [7, 8, 12, 9] as described in the next sections. Fifteen country names have been replaced with the naming notation used in the original dataset to perform the successful matching.

**Normalized Daily Spread Speed Estimation:** In this study, we analyzed the last two weeks of reported data from 14 March 2020 till 28 March 2020. To estimate the disease spread speed of each day in the two-week interval, we subtracted the reported number of new cases and fatalities on the previous day from the number of the current day and then divided this number to the Median reported number during the past two weeks.

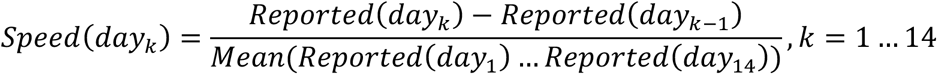

where *Reported()* is the number of Confirmed Cases or Fatalities reported in the dataset, *Mean()* is the Arithmetic Mean of its arguments. The above normalization procedure mitigates the problem of uneven speed of spread of COVID-19 in different geographical regions as it treats each country according to its outbreak “stage” and makes country statistics comparable to each other.

**Sparse Data Indicator Filtering:** As some of the data indicators in the merged dataset were found to contain a large number of missing values, which might affect further analysis, we excluded data indicators that contained more than 35% of missing values. After the sparse data indicator exclusion, the resulting dataset contained 130 data indicators.

### Correlation Analysis

To determine the relationship between the COVID-19 Spread Speed and other indicators, we applied Pearson’s product-moment Correlation [13] to 286 data samples (the countries and regions in the combined dataset) and 130 data indicators (whose data indicators that have remained after the Sparse Data Indicator Filtering step). We then filtered out all non-significant correlation values (α = 0.05) and presented the obtained results in the form of a correlation semi-matrix for visualization purposes. We list all the variables that have been incorporated into the correlation analysis in Multimedia Appendix 1. The Min, Max, Mean, Std. Deviation, Variance, Skewness, Kurtosis, and Overall Sum statistics are provided in Multimedia Appendix 2.

### Cluster Analysis

By adopting correlation analysis, we have determined multiple economic and medical system & health factors that are strongly and significantly correlated to the COVID-19 disease spread. We have also witnessed various governments and population behavioural traits possibly explaining the different scenarios of COVID-19 development around the world.

Even though these findings bring more light into the approaches that governments have adopted to mitigate the crisis, it is still unclear what the exact differences are in these approaches, as well as in the country profiles affected by the COVID-19 disease spread.

Aiming at answering this question, we have further adopted a Clustering Analysis technique [27] to study the groups of countries in our dataset by separating them based on the economic and medical system & health factors. In general, clustering analysis is a widely-adopted unsupervised machine learning technique allowing for automatic discovery of the groups of population samples in a multi-dimensional space of variables. Such groups could be then used for an in-depth understanding of the worldwide traits related to COVID-19 development, specifically with relation to various economic and health system factors. In this study, we have adopted the “X-Means” clustering algorithm, that has been reported to be effective in determining the number of clusters in the dataset without necessarily having a prior assumption on the number of clusters [28]. The X-Means clustering was applied with the bin value b = 1.0 and the cutoff factor c = 0.5, which were found empirically to prove a stable split of the data into clusters. Aiming at avoiding a potential bias in clustering results that could be introduced by various human decisions made when creating GHS index, the clustering has been performed in a reduced space excluding all GHS-related variables. In such a way, we are guaranteed that GHS has not impacted the clustering results, and therefore GHS could be used as a benchmark when evaluating the results of the analysis. Being guided by the value of the Bayesian Information Criterion (BIC), the X-Means clustering algorithm have determined the following four country-level clusters:

**Country Cluster 1:** Afghanistan, Angola, Bangladesh, Benin, Bhutan, Burkina Faso, Cambodia, Cameroon, Central African Republic, Chad, Côte D’Ivoire, Djibouti, Equatorial Guinea, Eritrea, Ethiopia, Gabon, Ghana, Guinea, Guinea-Bissau, Haiti, India, Indonesia, Kenya, Liberia, Madagascar, Mali, Mauritania, Mozambique, Namibia, Nepal, Niger, Nigeria, Pakistan, Philippines, Rwanda, Senegal, Somalia, Sri Lanka, Sudan, Tanzania, Timor-Leste, Togo, Uganda, Zambia, Zimbabwe.

**Country Cluster 2:** Albania, Algeria, Andorra, Antigua And Barbuda, Argentina, Armenia, Azerbaijan, Bahrain, Barbados, Belarus, Belize, Bolivia, Bosnia And Herzegovina, Brazil, Brunei, Bulgaria, Cabo Verde, Chile, Colombia, Costa Rica, Cuba, Cyprus, Dominica, Dominican Republic, Ecuador, Egypt, El Salvador, Fiji, Georgia, Grenada, Guatemala, Guyana, Holy See, Honduras, Iran, Iraq, Jamaica, Jordan, Kazakhstan, South Korea, Kuwait, Kyrgyzstan, Laos, Lebanon, Libya, Liechtenstein, Malaysia, Maldives, Mauritius, Mexico, Moldova, Mongolia, Montenegro, Morocco, Nicaragua, Oman, Panama, Papua New Guinea, Paraguay, Peru, Qatar, Romania, Saint Kitts And Nevis, Saint Lucia, Saint Vincent and The Grenadines, San Marino, Saudi Arabia, Serbia, Seychelles, Singapore, South Africa, Suriname, Syria, Thailand, Trinidad And Tobago, Tunisia, Turkey, Ukraine, United Arab Emirates, Uzbekistan, Venezuela;

**Country Cluster 3:** Australia (8 regions), Austria, Belgium, Canada (10 regions), Croatia, Czechia, Denmark, Denmark, Denmark, Estonia, Finland, France(10 regions), Germany, Greece, Hungary, Iceland, Ireland, Israel, Italy, Japan, Latvia, Lithuania, Luxembourg, Malta, Monaco, Netherlands (4 regions), New Zealand, North Macedonia, Norway, Poland, Portugal, Russia, Slovakia, Slovenia, Spain, Sweden, Switzerland, United Kingdom (7 regions), Uruguay, US (54 regions).

**Country Cluster 4:** China (33 regions).

We then treated each cluster assignment as independent variables and applied correlation analysis to uncover the statistical profiles for each of the clusters.

## Results

### Correlation Analysis

#### Correlation Visualization

**Multimedia Appendix 3:**
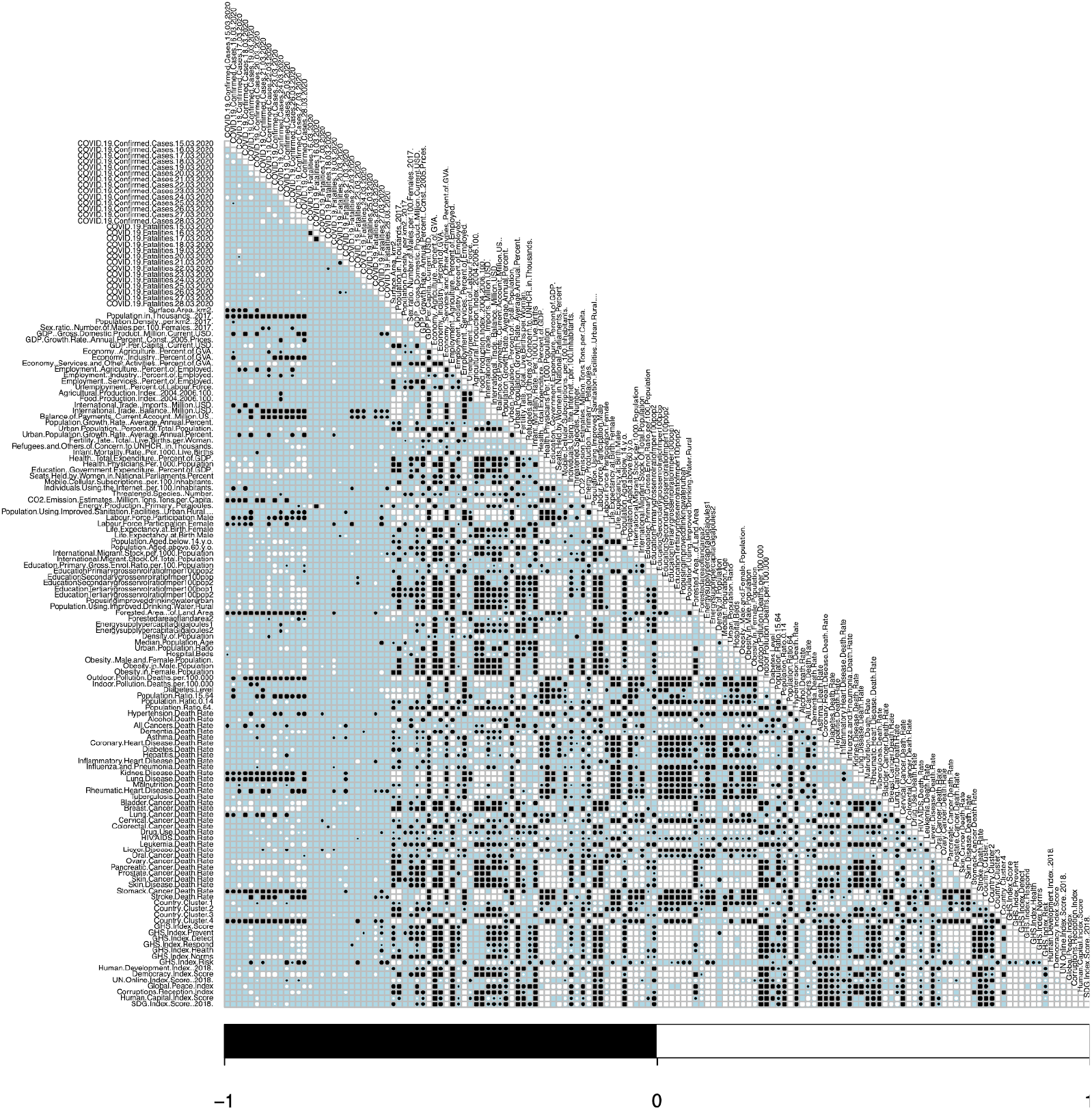
Pearson Product-Moment Correlation Between COVID-19 Spread Speed, Economics, and Health Factors.

For visualization purposes, we present the obtained correlation values in the form of a correlation semi-matrix (see Multimedia Appendix 3). In the Figure, white circles denote a positive correlation, while the black circles mean that the correlation is negative. The size of the circle is proportional to the correlation strength (the larger the circle - the stronger the correlation) and the absence of a circle in a cell means that there was no correlation found or the correlation is not significant.

#### Correlation Analysis of COVID-19 Confirmed Cases and Fatalities

From the first 28 lines of the correlation semi-matrix, it can be seen that there are several strong correlations between individual COVID-19 reported statistics. For example, it can be seen that there is a **strong negative correlation** between the **Fatality Speed on March 15 (Sunday) and March 16 (Monday)** as well as the **strong positive correlation** between **Fatality Speed on March 15 (Sunday) and March 17 (Tuesday)**. At the same time, a significant positive correlation was also found between several subsequent Confirmed Case Speed dates, such as 19 (Thursday), 20 (Friday), and 21 (Saturday) March 2020 and 26 (Thursday), 27 (Friday), and 28 (Saturday) March 2020.

Despite several single negative or positive correlations mentioned above, one might **not find many significant correlations** either between consequent measurements of the same metric (i.e. Confirmed Case Speed on Different Days) or between different COVID-19 metrics (i.e. Confirmed Case vs. Fatalities).

#### Correlation Analysis of Chronic Diseases and Health Factors

From the last 39 Chronic Disease and Health Factor indicators in the lower part of the correlation semi-matrix, it can be seen that there are multiple significant and consistent correlations of Chronic Disease Rates with Confirmed Case Speed measurements. For example, such indicators as **Skin Cancer** (91.7% 5-year survival rate [14]), **Prostate Cancer** (98.6% 5-year survival rate [14]), **Ovary Cancer** (46.5% 5-year survival rate [14]), **Breast Cancer** (89.7% 5-year survival rate [14]), and **Bladder Cancer** (77.3% 5-year survival rate [14]) Death Rates were found to be **significantly positively correlated** with the COVID-19 Spread Speed. In addition to the above findings, we would also like to highlight the **strong positive correlation** of **Obesity Rates (especially for Female demographics)** to COVID-19 Spread Speed. Furthermore, one can find that such variables as **Skin Disease Death Rate**, **Influenza and Pneumonia Death Rate**, **Diabetes Death Rate**, **Dementia Death Rate**, **Alcohol Death Rate** are also significantly correlated to COVID-19 Spread Speed. We would like to also highlight the significant positive correlations of most of the above factors to GHS Norm and GHS Health Indexes.

#### Correlation Analysis of Economic and Other Factors

From the correlation semi-matrix, it can be seen that six economic attributes are strongly positively correlated to the COVID-19 Spread Speed. First, we found a strong positive correlation between the **Number of Health Physicians Per 1000 Population, Health Total Expense (% of GDP)**, and **GDP Per Capita (in USD)** to the COVID-19 disease spread speed. Second, it can also be seen that **International Migrant Stock Per 1000 Population** and **International Migrant Stock % of Total Population** variables are also strongly and positively correlated to COVID-19 Spread Speed during the second week of the observed period (22 March 2020 - 28 March 2020). Third, one can notice a strong positive correlation of the **Services and Other Activities % of Gross Value Added (GVA)** measurement to COVID-19 disease spread. Finally, it can be seen that the GHS Detect Index variable is strongly positively correlated to at least five measurements of COVID-19 spread and two measurements of COVID-19 mortality towards the end of the observed 14-day interval. At the same time, it can also be observed that the GHS Democracy Index is strongly positively correlated to most of the COVID-19 spread and multiple COVID-19 mortality measures.

#### Negative and Insignificant Correlations, Mortality Rate Speed

An interesting observation is the strong **negative correlation** of the **Population in Thousands 2017** variable to the COVID-19 Spread Speed. At the same time, one can notice that the **Population Density Per km2 in 2017** does not exhibit any significant correlation with COVID-19. Furthermore, the reader could observe that such variables as **CO2 Emission Estimates (Million Tonnes Per Capita)** and **Forested Area Ratio** also exhibit a strong **negative correlation** to the COVID-19 Spread Speed. **Finally**, the **Estimates** metric is strongly **positively correlated** to the **Lung Cancer Death Rate**, which, in turn, is also **negatively correlated** to the **COVID19 Spread Speed**. Interestingly, such a relationship could not be observed from the data we have, except for the **only one negative correlation** of the **Lung Disease Death Rate** to the **COVID19 Fatality Speed on 23 March 2020**. Moreover, we would like to highlight the **International Trade Balance (Million USD)** and **Balance of Payment Current Account (Million USD)** are strongly **negatively correlated to the COVID-19 Fatality Speed** during the second week of the observed period (22 March 2020 - 28 March 2020).

#### Global Health Index (GHS) and Related Indexes Correlations

First, from the correlation semi-matrix, it can be seen that **all but one (GHS Index Risk) GHS Indexes are significantly positively correlated to each other**. Second, it can also be noticed that most of the GHS Index measurements are significantly correlated to COVID-19 disease spread speed when it comes to the end of the observed 14-day interval. The most significant positive correlations 8 and 9, were found for GHS Norms Index and GHS Democracy Index, respectively. The least number of correlations (two) were found for GHS Health Index.

### Cluster Analysis

As it was outlined in the Methods section, by applying X-Means clustering to the whole dataset, we have determined four country-level clusters and further applied correlation analysis treating each cluster assignment as an independent variable. The correlations between each cluster and other economic and health factors are visualized in the correlation semi-matrix in Multimedia Appendix 3.

#### Correlation Analysis of Country Cluster 1

From the correlation semi-matrix, it can be seen that the countries from the Cluster 1 are **positively correlated to the COVID-19 Confirmed Case Speed on 16, 20, and 23 March 2020**, while **negatively correlated on 25 March 2020**.

#### Correlation Analysis of Country Cluster 2

When looking at the correlation profile of the Country Cluster 2, a reader could immediately notice that the cluster is not associated with any significant COVID-19 correlations except for one positive correlation with COVID-19 Spread Speed on 17 March 2020. Furthermore, one can also find that **other positive correlations** of the cluster are **arguably weak**, having its spikes in **population growth** (see the significant positive correlation of Population Growth Rate Average Annual Percent variable to countries in Cluster 2), **Obesity** (see significant positive correlations of Obesity in Female Population and Obesity in Male Population variables to countries in Cluster 2) and **Diabetes** (see significant positive correlations of Diabetes Level and Diabetes Death Rate variables to countries in Cluster 2), various **heart-related diseases** (see significant positive correlations of Coronary Heart Disease Death Rate and Inflammatory Heart Disease Death Rate variables to countries in Cluster 2), and **reproduction system cancers** (see significant positive correlations of Cervical Cancer Death Rate and Prostate Cancer Death Rate variables to countries in Cluster 2). We would like to note that from the correlation semi-matrix it can be seen that the countries from the Cluster 2 are strongly negatively correlated to all GHS Index variables, except for the GHS Risk Index.

#### Correlation Analysis of Country Cluster 3

Country Cluster 3 is the largest and also the most diverse cluster that we have discovered as it includes most of the European Countries and all states of the US that have experienced spikes in COVID-19 cases over the past several weeks (see multiple significant positive correlations with COVID-19 spread speed and fatalities on the correlation plot).

From the correlation semi-matrix it can be seen that the countries from the cluster are **significantly positively correlated** to multiple factors associated with **modern developed economies**, such as higher **GDP Rate** (see See significant positive correlation of GDP Per Capita in USD variable to countries in Cluster 3), the involvement of the population in **Services Industry** (see significant positive correlations of Economy Services and Other Activities Percent of GVA and Employment Services Percent of Employed variables to countries in Cluster 3), high **Ratio of Urban Population** (see significant positive correlation of Urban Population Percent of Total Population variable to countries in Cluster 3), **Health System Maturity Level** (see significant positive correlations of Health Total Expenditure Percent of GDP, Health Physicians Per 1000 Population, Life Expectancy at Birth Female, and Life Expectancy at Birth Male variables to countries in Cluster 3), and solid **International Migrant Stocks** (see significant positive correlations of International Migrant Stock Per 1000 Population and International Migrant Stock Of Total Population variables to countries in Cluster 3).

At the same time, it can also be seen that countries from Cluster 3 exhibit a strong **positive correlation with population ageing and its associated diseases** (see significant positive correlations of Median Population Age, Population Ratio 64+ Years Old, and Dementia Death Rate variables to countries in Cluster 3), various types of **cancers** (see significant positive correlations of Bladder Cancer Death Rate, Breast Cancer Death Rate, Colo-rectal Cancer Death Rate, Leukemia Death Rate, Ovary Cancer Death Rate, Pancreatic Cancer Death Rate, and Skin Cancer Death Rate variables to countries in Cluster 3) and, correspondingly, **urban population-linked chronic diseases** (see significant positive correlations of Obesity Male and Female Population, Obesity in Female Population, Obesity in Male Population, Drug Use Death Rate, Skin Disease Death Rate, and Alcohol Death Rate variables to countries in Cluster 3).

Finally, from the correlation semi-matrix, it can be seen that countries from Cluster 3 are strongly positively correlated to almost all GHS Index variables.

#### Correlation Analysis of Country Cluster 4

As China is the only country in Cluster 4 and its economic, population, and, for example, pollution statistics are commonly known, we will omit some strongly positively correlated indicators in this work. Examples of such **strong positive** correlations are **Lung Disease Death Rate**, **Stomach Cancer Death Rate**, **Malnutrition Death Rate Rheumatic**, and **Heart Disease Death Rate**, that is also strongly **negatively correlated to the COVID-19 Spread Speed**.

## Discussions

In the previous section, we discovered that multiple economic factors exhibit a strong relationship to the chronic diseases across the globe, and, therefore, it would be reasonable to hypothesise that they can be utilized to characterize the profiles of these countries with relation to the economic development stage, and ultimately, COVID-19 Spread Speed.

To gain further insights into the relationship between such economic factors and the COVID-19 disease spread, in this section we will discuss the possible reasons for the discovered relationships.

### Correlation Analysis

#### Correlation Analysis of COVID-19 Confirmed Cases and Fatalities

The negative correlation between the Fatality Speed on Sunday and Monday, as well as the strong positive correlation between Fatality Speed on Sunday and Tuesday, could possibly suggest a reporting time-lag on weekends. At the same time, the discovered correlation sequences towards the end of the week could be explained by the testing capacity of the medical institutions entailing the situation when test results from the beginning of the week were received only towards the end of the week. As this study does not aim at a detailed analysis of the longitudinal properties of the COVID-19 measurement and test procedures [36], we would only like to **highlight the importance and the influence of time-related measurement and testing arrangement aspects** as well as to recommend future research along this direction.

The absence of significant correlations neither between consequent measurements of the same disease spread metric (i.e. Confirmed Case Speed on Different Days) nor between different COVID-19 metrics (i.e. Confirmed Case vs. Fatalities) reveals that there is no strong linear relation between Confirmed Case Speed and Fatality Speed within the 14 day-interval and in the space of independent variables that have been analyzed in this study. Therefore, **we recommend the utilization of additional data sources, such as hospital capacity, testing volume, internal government regulations and policies border closure etc**. for gaining a deeper insight into the actual relationship between COVID-19 Infection and Fatality trends.

#### Correlation Analysis of Chronic Diseases and Health Factors

In order to explain the discovered significant positive correlations between Chronic Disease Rates and Confirmed Case Speed, it is necessary to consider the factors related to country-level chronic disease data indicators.

First, it was previously reported in the literature [17] that COVID-19 development and consequences might be directly related to the overall health status of the population, especially with regards to the existing pre-conditions affecting the human immune system. Such pre-conditions could further entail various fatal complications, such as Cytokine Storm, and, ultimately, affect the countries’ COVID-19 Confirmed Case Speed and Fatality Speed statistics. In this study, we could have also witnessed such relationships in our results. Specifically, the correlation semi-matrix reveals several potential indicators of poorer population immunity in a large portion of analyzed countries, which was reflected in our correlation analysis results by the significant positive correlation of e.g. Skin Disease Death Rate, Influenza and Pneumonia Death Rate, Diabetes Death Rate, Dementia Death Rate, and Alcohol Death Rate to COVID-19 Spread Speed.

Second, a possible indicator of the health system weaknesses could be found among significant positive correlations between COVID-19 Spread Speed and various types of Cancers Death rates, especially those cancers with an average higher survival rates in the developed world. In particular, it is reasonable to assume that the countries exhibiting higher death rates for such “high-survival cancers’’ might experience overall difficulties in proper and timely patient treatment. When facing COVID-19 pandemic, such countries might not be always well prepared for proper patient isolation and treatment as well, which is essential for COVID-19 disease spread control [15,37,38]. Correspondingly, in such countries, the COVID-19 Spread Speed could be higher entailing the above-reported significant positive correlation [16].

To summarize, in this section **we have discovered two potential traits** affecting the speed of COVID-19 disease spread. The first finding suggests that it could be possible that the **significant correlations between chronic diseases death rates** and COVID-19 Spread Speed, especially those diseases that are **tightly knit to the human immune system**, could **reflect the overall country medical system & population health status** and, therefore, predisposition to infection and complication of COVID-19. Even though operationally, the more developed economies could, arguably, respond faster to COVID-19 outbreak, **such populations might be also more affected by various urban-living factors** [18, 19], that, in turn, could **entail COVID-19 health predispositions and skewed disease spread statistics**. The second finding highlights the possible causality between the ability of health systems to control the rapid infections disease outbreaks efficiently and the speed of COVID-19 spread. This conclusion is also supported by the negative correlations found, for example, between the GHS Health System and Compliance Index and the Mortality rates with such diseases as Influenza and Pneumonia.

As a final note, we would like to state that in this study we are not attempting to compare various types of Cancers, Chronic Diseases, and COVID-19 disease directly, as they are known to be very different in terms of their cause, progression, and detection/treatment principles. Instead, we are aiming to treat them as independent statistical variables that, as have been shown above, could both reflect and, potentially, predict the evolution pace of the COVID-19 disease.

#### Correlation Analysis of Economic and Other Factors

When it comes to explaining the statistical relationships between COVID-19 spread and various Factors describing Economic systems, it is reasonable to start from the basic criteria related to economic strength, especially with relation to the basic health metrics. Specifically, the strong positive correlation of Health Physicians, Health Total Expense, and GDP Per Capita variables could be attributed to the higher ability of the countries with stronger health systems in performing timely patient assessment, diagnosis, and disease reporting. In contrast, countries with weakly-subsidized health systems, many, especially asymptomatic [20], **COVID-19 cases could remain unreported bringing the COVID-19 confirmed case statistics down** and entailing the inverse correlation traits that we have discovered from the dataset. The finding aligns well with the significant correlation discovered between COVID-19 disease spread and the GHS Detect Index. Particularly, the correlations suggest that **economies with average higher disease detection ability might have reported the higher COVID-19 increase rates**, as opposed to to the countries where the virus might have been spreading in the community but left undetected and unreported.

In the cases of International Migrant Stock Per 1000 Population and International Migrant Stock % of Total Population variables, the discovered strong positive correlations to COVID-19 Spread Speed could be possibly explained by the higher rates of imported cases in countries with larger proportions of the migrant population who often travel abroad or within the country [39] for business and personal purposes. Interestingly, both variables exhibit a strong positive correlation during the second week of the observed period (22 March 2020 - 28 March 2020), which could be potentially explained by the travel restrictions imposed by the governments during that week, resulting in the situation when many migrants were rushing to return back to their countries of residence prior to border closures [21, 22]. Finally, the strong positive correlation of the Services and Other Activities *%* of Gross Value Added (GVA) measurement can be attributed to the more intense human interaction rates in countries with larger populations involved in the service sector of economics, making the risk of COVID-19 infection higher [23].

Summarizing the above, again, we would like to **highlight the importance of incorporation in the analytics of the data related to the particular lockdown policies enforced by the governments attempting to cess the COVID-19 spread rapidly**. For example, more centralized governments, such as China, Singapore and Vietnam, could be able to implement effective lockdowns faster as compared to less centralized political systems. This facilitates tighter control and faster relief from the disease, while, possibly, temporarily sidestepping human rights and privacy concerns, which reflect similar social concerns during the SARS outbreak [45]. Opposingly, the European Union countries took much longer time to respond to the COVID-19 with the necessary population restriction measures and those measures were found to be less effective in terms of population compliance [40]. This might be due to countries operating with different political groups, which have been found to hinder a unified or equitable approach when dealing with large-scale public health concerns [46]. Such **relation of the country’s political system and the speed of effective response is also supported by the findings of this study**. Specifically, from the correlation semi-matrix, it can be seen that the GHS democracy index is correlated to the majority of the COVID-19 spread measures as well as to many COVID-19 mortality measures during the observed 14-day interval.

#### Negative and Insignificant Correlations, Mortality Rate Speed

When it comes to explaining the Negative and Insignificant Correlations as well as the correlations with Mortality Rate Speed, there are several interesting data relationships that can be observed. For example, the strong negative correlation of the population size along with the absence of a significant correlation of population density with COVID-19 could be possibly explained by the inexistence of a linear relationship between these factors and COVID-19 Spread Speed. As it was previously reported [24] and was also observed in this study, the cultural and behavioural factors, such as **human interaction habits**, or government-regulated factors and measure enforcement viability, such as **social distancing enforcement measures could be of a much higher influence on the ability of the country government to manage the COVID-19 disease outbreak**.

Furthermore, the strong negative correlation of the Forested Area Ratio to the disease spread speed can be hypothesized by the natural geographical sparsity of population introduced by the forested landscape and entailing a limited inter-human interaction.

At the same time, the strong negative correlation of the CO2 Emission variables requires additional clarifications. One possible explanation arises by also taking into consideration the strong positive correlation of the metric to the Lung Cancer Death Rate, which, in turn, is also negatively correlated to the COVID19 Spread Speed. Precisely, taking into consideration that the two metrics might not be related directly to the disease spread speed (as COVID-19 disease gets “…transmitted between people through close contact and droplets” [25] and, thus, more depends on the inter-human close contacts), it is then reasonable to assume that the two variables could also be positively correlated to the COVID-19 Fatality Speed as we could expect more patients with lung preconditions in the countries with more polluted environments. However, such a relationship could not be observed from the data available in this study, and the only one negative correlation of the Lung Disease Death Rate to the COVID19 Fatality Speed on 23 March 2020 could be due to the reporting bias.

The strong negative correlation of COVID-19 fatality to the International Trade Balance and Balance of Payment variables observed during the second week of the data (22 March 2020 - 28 March 2020) can be possibly explained as that both variables reflect the countries’ ability to manage the spiking COVID-19 disease outbreak: countries with stronger economies and medical equipment reserve might be able to provide patients with necessary care when being pressured by the high daily case numbers, as compared to the economies experiencing a shortage of resources.

Overall, the high sparsity of the correlation semi-matrix regarding the COVID-19 Fatality Speed metrics might possibly suggest that, **at the observed time interval the COVID-19 data on Fatality Rate might have been not sufficient for making conclusive observations on the inter-variable relationships and further research on more recent data is necessary**.

#### Global Health Index (GHS) and Related Indexes Correlations

Last but not least, in this study, we would like to discuss the GHS Metrics and their relation and applicability to the COVID-19 disease outbreak.

First, from the strong significant mutual correlation between GHS indexes, we could immediately notice a possible insufficient comprehensiveness of the metric for measuring infections disease outbreaks in a multi-factor health and economics environment. Specifically, as it is stated by the authors [40], the index has been constructed based on “…140 questions, organized across 6 categories, 34 indicators, and 85 sub-indicators to assess a country’s capability to prevent and mitigate epidemics and pandemics “. Judging from the correlation analysis results, it can be seen that the **categories indeed are highly correlated to each other and therefore might be redundant**.

Second, when considering the specific GHS Index components, such as **GHS Norm Index**, they **show the ability to reflect the COVID-19 disease progression** most of the time in the observed interval, and therefore could potentially be a **better indicator for COVID-19 progression forecasting** (9 Significant correlations with COVID-19 measurements our of 28 possible) **as compared to the combined GHS Index** (6 significant correlations with COVID-19 measurements out of 28 possible). The finding highlights the importance of considering the “Compliance With International Norms” factors for COVID-19 pandemic mitigation and forecast, namely: cross-border agreements on public health emergency response; international commitments; completion and publication of WHO JEE and the World Organization for Animal Health (OIE) Performance of Veterinary Services (PVS) Pathway assessments; financing; and commitment to sharing of genetic and biological data and specimens [40].

Finally, we would like to highlight that the most **COVID-correlated variable was found to be GHS Democracy Index**, **again suggesting that countries’ political systems play a crucial role defining the pace** in which countries could adopt and enforce disease control and prevention measures and, therefore, mitigate the COVID-19 progression. On the contrary, the **GHS Health Index was found to be least correlated to COVID-19**, revealing that such metrics as health capacity in medical institutions; medical countermeasures and personnel deployment; healthcare access; communications with healthcare workers during a public health emergency; infection control practices and availability of equipment; and capacity to test and approve new countermeasures might not be a strong predictive indicator of COVID-19 pandemic development.

To conclude this section, some of the GHS-adopted indicators were found to be of a tight relation to COVID-19 disease development and, therefore, could potentially serve as a source of the disease prediction and prevention. However, **GHS index model was also found to be simplistic and redundant** when being applied to COVID-19 data, highlighting the importance of only two variables (GHS Norm Index and GHS Democracy Score) for COVID-19 outbreak analysis. We, therefore, **recommend the adoption of other public policy and political system-related measures as well as more sophisticated non-linear models when attempting to analyze and predict the COVID-19 pandemic development**.

### Cluster Analysis

In the previous sections, we have discussed multiple economic and health factors that are strongly and significantly correlated to the COVID-19 disease spread. We have also witnessed various governments and population behavioral traits possibly explaining the different scenarios of COVID-19 development around the world. Even though the above-discovered findings bring more light into the way that governments could adopt to mitigate the crisis, it is still unclear what the exact differences in the country profiles affected by the COVID-19 disease spread and how the country grouping can be explained. Below, we provide such explanations for most of the discovered correlation relationships.

#### Correlation Analysis of Country Cluster 1 Correlations

The correlations of the Cluster 1 described in the Results section could characterize the countries from the cluster as belonging to the category of developing world, which could also be observed from the cluster-country member list provided earlier. Therefore, the non-consistent correlations with the COVID-19 Confirmed Case Speed (three significant positive correlations and one significant negative correlation), could then be explained by possible testing and reporting issues that frequently occur when facing world-scale disease outbreaks [29]. Given the limited available data in our COVID-19 dataset regarding COVID-19 reporting procedures in different countries, in this work, we would like to **highlight a possible under-reporting issue for the developing world** [34], implying not just biased statistics of the datasets used for COVID-19 analysis, but also possible wrong perception and underestimation of the pandemic impact on people lives and world economies, unavoidably entailing higher infection/mortality rates and crisis escalation. Consequently, we suggest further in-depth research towards COVID-19 spread characteristics in the developing countries taking into consideration alternative measures of disease progression evaluation in parallel with official statistics [44].

#### Correlation Analysis of Country Cluster 2 Correlations

When analyzing the correlation profile of the Country Cluster 2, a reader could immediately notice that the cluster exhibits weak positive correlations having its spikes in population growth, Obesity, and Diabetes, various heart-related diseases, and reproduction system cancers. From the observed relationships, we can acknowledge that the countries in the cluster can be characterized by the population overweight, and correspondingly, heart [30] and reproductive cancer problems [31]. Furthermore, it also can be seen that the cluster is not associated with any significant COVID-19 correlations except for one positive correlation with COVID-19 Spread Speed on 17 March 2020. A possible reason for the correlation absence could be the “noise” in the data that is introduced by the operational challenges that the countries experience when measuring and reporting the COVID-19 disease cases. For example, discussing Brazil (a member of the Cluster 2), Cost Ribeiro et. al. [34], have witnessed that “…the numbers reported by the Brazilian government should be far from the real situations.” and suggested that “…the confirmed number of cases must be multiplied by a factor of 7.7 to obtain the actual number of infected patients in hospital conditions.” In other countries, for example in the US, the multiplier was reported to be even higher, reaching the value of 8 [35]. With such a high reporting bias, it is reasonable to assume that the **COVID-19 data about the countries from Cluster 2 could possibly be heavily biased restricting the statistical methods such as Correlation Analysis to discover significant relationships between COVID-19 and the cluster they belong to**. The latter observation is also indirectly supported by the discovered negative correlations of Cluster 2 countries to all Global Health Index variables except for the Risk Factor Index. In particular, the countries that are, conventionally, evaluated as to be belonging to a “higher risk” group are, surprisingly, exhibiting no correlation to COVID-19 progression points out, once more, that there is a possible bias in evaluation introduced by the quality of COVID-19 reporting.

#### Correlation Analysis of Country Cluster 3 Correlations

From the correlations highlighted in the Results, it can be seen that the countries from the cluster are significantly positively correlated to multiple factors associated with typical modern developed economies. At the same time, it can also be seen that these countries exhibit a strong positive correlation with ageing population and its associated diseases. Taking into consideration these two traits and the multiple observed correlations of Cluster 3 with COVID-19 variables, we could hypothesize that t**he Cluster 3 members are mostly developed economies and that their populations might be also initially predisposed to COVID-19 infection entailing multiple strong positive correlations with COVID-19 disease spread and fatality rates**. The latter assumption raises from the two known COVID-19 risk factors that are also to be found related to the countries from Cluster 3, namely older population demographics [32] and existing pre-conditions that could lead to, for example, Cytokine Storm [17] or other highly-lethal COVID-19 complications. Finally, it can be seen that the countries in the Cluster 3 are strongly correlated with almost all GHS Indexes, suggesting that **GHS indexing system, indeed tends to score developed economies higher and might not be a suitable metric when predicting disease outbreaks in the light of disease underreporting, political system differences, and the gaps of development between different economic systems**.

#### Correlation Analysis of Country Cluster 4 Correlations and Possible Limitations

When talking about the Cluster 4, which solely consists of the data from China, we would like to bring the readers’ attention to the possible bias in some of the conclusions that we have drawn from the data in this study. By drawing a parallel between the observed positive correlations of China-specific chronic diseases as well as the negative correlation with the COVID-19 spread speed, readers could conclude that **data from China might affect the overall analysis results in relation to the shift of the disease development timeline between China and other countries**. We, therefore, recommend considering such potential time biases in future quantitative research, as such data points might significantly affect the prediction analysis results when being analyzed jointly with other indicators. Furthermore, we would also like to highlight the importance of the proper alignment and synchronization of the data that comes from the regions with large territories and specific disease development timelines. Lastly, we would like to reiterate the possible shortcomings of our-operated dataset related to the underreporting issues introduced by multiple countries. More precisely, in some scenarios, the low number of reported COVID-19 cases could be explained by underreporting while in others - by successfully employed COVID-19 control measures. The former might mislead governments when evaluating the performance of the latter, and inverse, the latter might not have proper feedback on their COVID-19 control measures when benchmarking themselves against the former. Not to say that all the above could potentially bias the statistical analysis results and the prediction models that could attempt to forecast COVID-19 progression based on various economic and health system factors.

## Conclusions

In this preliminary research, we have identified the possible underreporting issue of COVID-19 disease. We have also highlighted four scenarios of COVID-19 development determined from the country-level cluster analysis study and outlined the shortcoming of the existing disease measurement approaches, such as Global Health Index (GHS) scoring. We believe that our work would encourage further in-depth quantitative research along the direction as well as would be of support to public policy development when addressing the COVID-19 crisis worldwide.

Specifically, we would like to highlight the following key findings and recommendations that have been identified in this research:

- **Longitudinal COVID-19 data on cases and mortality rates alone might not be sufficient for forecasting/predicting COVID-19** situations across the world and the utilization of additional Economic and Health System data sources.
- **Population immunity and urban-living factors could serve as statistical indicators** reflecting the predisposition of countries to rapid infectious disease spread.
- **Reporting is a crucial factor** in understanding COVID-19 evolution. For the less developed economies, many COVID-19 might have been left unreported or misclassified, while for the more developed economies, the COVID-19 disease progression speed might have been reported higher due to well-organized testing and reporting. Overall **the testing ability and the number of registered cases factors must be always considered together** for making conclusive observations on COVID-19 progression.
- The **political system was found to be another crucial factor affecting the successful implementation of COVID-19 preventive measures**, such as lockdowns and border closures. **GHS Democracy and Norm Indexes were found to be of a high relation to COVID-19 development**, **GHS Health Index was found to be redundant and weakly related to COVID-19 progression** during the observed 14-day time interval.
- **GHS index model was found to be simplistic and redundant when measuring and forecasting COVID-19 spread and new more comprehensive** (data-vice and architecture-ice models) **models are necessary** to be developed.

## Future Work

To encourage future studies on predictive COVID-19 analytics, in this work we implemented a preliminary test of Regression analysis by applying Linear Regression Model onto our datasets and fitting the model for predicting the COVID-19 Spread Speed during the last day of the observed 14-days interval. As many of the employed variables were found to be correlated to each other, in order to achieve a balanced regression fitting, prior to running regression fitting we have also applied Principal Component Analysis (PCA) [43] that have resulted in projecting the data in a new space consisting of 132 principal components (determined automatically by PCA preserving 100% of variance). The results of the test are presented in Multimedia Appendix 4. From the results, it could be seen that the modulo values of at least 53 regression coefficients were found to be greater than 0.01 (p < 0.05), suggesting the applicability and the high potential of using our dataset in COVID-19 prediction task.

In future works, we are aiming at establishing automotive Machine Learning and Statistical frameworks that would be attempting to predict the future development of COVID-19 disease based on our COVID-19 dataset. We will be also extending the dataset with more dynamic and comprehensive data, such as medical resource availability, government-imposed control measures, and culture-driven aspects.

## Data Availability

The data that support the findings of this study are openly available: http://covid19.somin.ai/

## Acknowledgements

Aleksandr Farseev and Yu-Yi Chu-Farseeva conceived of the presented idea. Aleksandr Farseev and Qi Yang developed the theory and performed the data analysis. Yu-Yi Chu-Farseeva and Daron Benjamin Loo verified the analytical methods. Yu-Yi Chu-Farseeva encouraged Aleksandr Farseev to investigate population health-related aspects of the COVID-19 Disease Spread and Fatalities Speed and supervised the findings of this work. All authors discussed the results and contributed to the final manuscript.

## Conflicts of Interest

We declare no competing interests.

Abbreviations
COVID-19: Coronavirus disease 2019
GDP: Gross Domestic Product
GVA: Gross Value Added

## Multimedia Appendix 1

Variables Participated in Correlation Analysis

[XLSX file (Microsoft Excel), 53kB]

## Multimedia Appendix 2

Table of Statistics

[XLSX file (Microsoft Excel), 24kB]

## Multimedia Appendix 3

Pearson Product-Moment Correlation Between COVID-19 Spread Speed, Economics, and Health Factors

[PDF file, 124 kB]

## Multimedia Appendix 4

Regression Coefficients

[XLSX file (Microsoft Excel), 18kB]

## Multimedia Appendix 5

Pearson Product-Moment Correlation Between COVID-19 Spread Speed, Economics, and Health Factors

[XLSX file, 307kB]

## References

1. Y. Bai, L. Yao, T. Wei, F. Tian, D.-Y. Jin, L. Chen, M. Wang, Presumed asymptomatic carrier transmission of covid-19. Jama. PMID: 32083643

2. Z. Wu, J. M. McGoogan, Characteristics of and important lessons from the coronavirus disease 2019 (covid-19) outbreak in China: summary of a report of 72314 cases from the Chinese center for disease control and prevention. Jama. PMID: 32091533

3. Y. Ji, Z. Ma, M. P. Peppelenbosch, Q. Pan, Potential association between covid-19 mortality and health-care resource availability. The Lancet Global Health. PMID: 32109372

4. Z. Chen, Q. Zhang, Y. Lu, Z. Guo, X. Zhang, W. Zhang, C. Guo, C. Liao, Q. Li, X. Han, et al., Distribution of the covid-19 epidemic and correlation with population emigration from Wuhan, China.. Chinese medical journal. PMID: 32118644

5. M. Chinazzi, J. T. Davis, M. Ajelli, C. Gioannini, M. Litvinova, S. Merler, A. P. y Piontti, K. Mu, L. Rossi, K. Sun, et al., The effect of travel restrictions on the spread of the 2019 novel coronavirus (covid-19) outbreak, Science. PMID: 32144116

6. Covid19 global forecasting (week 2): forecast daily covid-19 spread in regions around world, https://www.kaggle.com/c/covid19-global-forecasting-week-2/data, accessed: 2020-03-29.

7. Country statistics – undata dataset, https://www.kaggle.com/sudalairajkumar/undata-country-profiles, accessed: 2020-03-29.

8. Pollution by country for covid19 analysist, https://www.kaggle.com/brandonhoeksema/pollution-by-country-for-covid19-analysis, accessed: 2020-03-29.

9. World life expectancy database, https://www.worldlifeexpectancy.com/, accessed: 2020-03-29.

10. Covid-19 combined dataset (somin.ai), https://covid19.somin.ai/, accessed: 2020-03-29.

11. S. A. Lauer, K. H. Grantz, Q. Bi, F. K. Jones, Q. Zheng, H. R. Meredith, A. S. Azman, N. G. Reich, J. Lessler, The incubation period of coronavirus disease 2019 (covid-19) from publicly reported confirmed cases: estimation and application. Annals of internal medicine. PMID: 32150748

12. The world bank data, https://data.worldbank.org/indicator/SP.POP.0014.TO.ZS, accessed: 2020-03-29.

13. D. Freedman, R. Pisani, R. Purves, Statistics (international student edition), Pisani, R. Purves, 4th edn. WW Norton & Company, New York.

14. Eer cancer statistics review, 1975-2013, https://seer.cancer.gov/archive/csr/1975_2013/, accessed: 2020-03-30.

15. J. Hellewell, S. Abbott, A. Gimma, N. I. Bosse, C. I. Jarvis, T. W. Russell, J. D. Munday, A. J. Kucharski, W. J. Edmunds, F. Sun, et al., Feasibility of controlling covid-19 outbreaks by isolation of cases and contacts. The Lancet Global Health PMID: 32119825

16. J. C. Wells, A. A. Marphatia, T. J. Cole, D. McCoy, Associations of economic and gender inequality with global obesity prevalence: understanding the female excess. Social science & medicine 75 (3) (2012) 482-490. PMID: 22580078

17. P. Mehta, D. F. McAuley, M. Brown, E. Sanchez, R. S. Tattersall, J. J. Manson, Covid-19: consider cytokine storm syndromes and immunosuppression. The Lancet. PMID: 32192578

18. V. L. Feigin, C. M. Lawes, D. A. Bennett, C. S. Anderson, Stroke epidemiology: a review of population-based studies of incidence, prevalence, and case-fatality in the late 20th century, The Lancet Neurology 2003;2(1):43-53 PMID: 12849300

19. M. Pereira, N. Lunet, A. Azevedo, H. Barros, Differences in prevalence, awareness, treatment and control of hypertension between developing and developed countries, Journal of hypertension. 2009;27(5);963–975 PMID: 19402221

20. Z. Hu, C. Song, C. Xu, G. Jin, Y. Chen, X. Xu, H. Ma, W. Chen, Y. Lin, Y. Zheng, et al., Clinical characteristics of 24 asymptomatic infections with covid-19 screened among close contacts in Nanjing, China, Science China Life Sciences (2020) 1-6 PMID: 32146694

21. Coronavirus: Travel Restrictions, border shutdowns by country, https://www.aljazeera.com/news/2020/03/coronavirus-travel-restrictions-border-shutdowns-country-200318091505922.html, accessed: 2020-04-04.

22. W. H. Organization, et al., Coronavirus disease 2019 (covid-19): situation report, 67.

23. J. Hilton, M. J. Keeling, Estimation of country-level basic reproductive ratios for novel coronavirus (covid-19) using synthetic contact matrices, medRxiv.

24. S. Merler, M. Ajelli, L. Fumanelli, M. F. Gomes, A. P. y Piontti, L. Rossi, D. L. Chao, I. M. Longini Jr, M. E. Halloran, A. Vespignani, Spatiotemporal spread of the 2014 outbreak of ebola virus disease in liberia and the effectiveness of non-pharmaceutical interventions: a computational modelling analysis, The Lancet Infectious Diseases 2015;15(2):204–211 PMID: 25575618

25. W. H. Organization, et al., Rational use of personal protective equipment for coronavirus disease (covid-19): interim guidance, 27 February 2020, Tech. rep., World Health Organization (2020).

26. T. Beck, Financial development and international trade: Is there a link?, Journal of international Economics 2002;57(1):107–131.

27. X. Wu, S. Hu, A. B. Kwaku, Q. Li, K. Luo, Y. Zhou, H. Tan, Spatio-temporal clustering analysis and its determinants of hand, foot and mouth disease in Hunan, china, 2009-2015, BMC infectious diseases 2017;17(1):645. PMID: 28946852

28. D. Pelleg, A. W. Moore, et al., X-means: Extending k-means with efficient estimation of the number of clusters. In Proceedings of the 17th International Conf. on Machine Learning, 2000;1:727–734.

29. L. Bakalikwira, J. Bananuka, T. Kaawaase Kigongo, D. Musimenta, V. Mukyala, Accountability in the public health care systems: A developing economy perspective, Cogent Business & Management 2017;4(1).

30. A. Keys, Obesity and heart disease, Journal of Chronic Diseases 1955;1(4)

31. N. M. Maruthur, S. D. Bolen, F. L. Brancati, J. M. Clark, The association of obesity and cervical cancer screening: a systematic review and metanalysis, Obesity 2009;17(2):375–381.

32. Q. Ruan, K. Yang, W. Wang, L. Jiang, J. Song, Clinical predictors of mortality due to covid-19 based on an analysis of data of 150 patients from Wuhan, China, Intensive care medicine 2020:1(3). PMID: 32253449

33. Kaggle, a subsidiary of Google LLC, is an online community of data scientists and machine learning practitioners. https://www.kaggle.com/

34. L. C. Ribeiro, A. T. Bernardes, Estimate of underreporting of COVID-19 in Brazil by Acute Respiratory Syndrome hospitalization reports, No 010, Notas Técnicas Cedeplar-UFMG, Cedeplar, Universidade Federal de Minas Gerais, https://EconPapers.repec.org/RePEc:cdp:tecnot:tn010.(2020)

35. Laboratory-Confirmed COVID-19-Associated Hospitalizations, preliminary cumulative rates as of May 23, 2020. https://gis.cdc.gov/grasp/COVIDNet/COVID19_3.html

36. Guest JL, Del Rio C, Sanchez T. The Three Steps Needed to End the COVID-19 Pandemic: Bold Public Health Leadership, Rapid Innovations, and Courageous Political Will. JMIR Public Health Surveill. 2020;6(2):e19043 PMID:32240972

37. Gilbert, M., Pullano, G., Pinotti, F., Valdano, E., Poletto, C., Boëlle, P. Y., … & Gutierrez, B. (2020). Preparedness and vulnerability of African countries against importations of COVID-19: a modelling study. The Lancet, 395(10227), 871–877. PMID:32087820

38. Rodriguez-Morales AJ, Gallego V, Escalera-Antezana JP, et al. COVID-19 in Latin America: The implications of the first confirmed case in Brazil [published online ahead of print, 2020 Feb 29]. Travel Med Infect Dis. 2020;101613. doi:10.1016/j.tmaid.2020.101613 PMID:32126292

39. Jia JS, Lu X, Yuan Y, Xu G, Jia J, Christakis NA. Population flow drives spatio-temporal distribution of COVID-19 in China [published online ahead of print, 2020 Apr 29]. Nature. 2020;10.1038/s41586-020-2284-y. doi:10.1038/s41586-020-2284-y PMID:32349120

40. Goniewicz, K.; Khorram-Manesh, A.; Hertelendy, A.J.; Goniewicz, M.; Naylor, K.; Burkle, F.M., Jr. Current Response and Management Decisions of the European Union to the COVID-19 Outbreak: A Review. Sustainability 2020, 12, 3838.

41. Cameron, E. E., Nuzzo, J. B., & Bell, J. A.(2019). Global health security index: Building collective action and accountability. Baltimore, MD: Johns Hopkins, Bloomberg School of Public Health. Retrieved from https://www.ghsindex.org/wp-content/uploads/2019/10/2019-Global-Health-Security-Index.pdf.

42. Andersen, Martin, Early Evidence on Social Distancing in Response to COVID-19 in the United States (April 6, 2020). Social Science Research Network. http://dx.doi.org/10.2139/ssrn.3569368

43. Wold, S., Esbensen, K., & Geladi, P. (1987). Principal component analysis. Chemometrics and intelligent laboratory systems, 2(1-3), 37–52.

44. Remuzzi, A., & Remuzzi, G. (2020). COVID-19 and Italy: what next?. The Lancet.

45. Teo, P., Yeoh, B. S., & Ong, S. N. (2005). SARS in Singapore: surveillance strategies in a globalising city. Health Policy, 72(3), 279–291.

46. Hicken, A., Kollman, K., & Simmons, J. W. (2016). Party system nationalization and the provision of public health services. Political Science Research and Methods, 4(3), 573–594.

